# The Impact of Doxycycline Post-Exposure Prophylaxis on Antibiotic Use at a Boston Sexual Health Clinic

**DOI:** 10.64898/2026.08.04.26359130

**Authors:** Rachel Mittelstaedt, David Helekal, Madeleine C. Kline, Kirstin I. Oliveira Roster, Gregory K. Robbins, Kevin L. Ard, Yonatan H. Grad

## Abstract

**Background:** Doxycycline post-exposure prophylaxis (doxy-PEP) reduces the incidence of bacterial sexually transmitted infections (STIs) among men who have sex with men and transgender women (MSMTW), but it may select for antimicrobial resistance (AMR). AMR development will depend in part how doxy-PEP changes rates of antibiotic use.

**Methods:** We conducted a retrospective electronic medical record review of antibiotic prescriptions received by patients at the Massachusetts General Hospital Sexual Health Clinic from January 1, 2023, to December 27, 2025. Using a Bayesian negative binomial regression, we assessed the direct, indirect, and combined effects of doxy-PEP implementation on antibiotic prescription rates among doxy-PEP-eligible MSMTW who were receiving HIV pre-exposure prophylaxis.

**Results:** Controlling for direct effects, the cohort’s total antibiotic prescription rate decreased by 10% (0.90, 95% CI 0.87 – 0.94) for every 100 doxy-PEP starts. Doxy-PEP users were prescribed antibiotics at double the rate predicted in the absence of doxy-PEP implementation, and, controlling for indirect effects, received 3.40 (95% CI 2.95 – 3.91) times the antibiotic prescriptions of patients not using doxy-PEP. Doxy-PEP non-users were prescribed antibiotics at less than half the rate predicted in the absence of doxy-PEP. The full cohort’s overall antibiotic prescription rate increased by 1.4 times after doxy-PEP implementation.

**Conclusions:** Individuals who are taking doxy-PEP have higher antibiotic prescription rates, increasing selection for antibiotic-resistant bacteria in these individuals. However, doxy-PEP-driven decreases in the overall incidence of bacterial STIs have the potential to decrease selective pressure for resistant organisms in those not using doxy-PEP.

## Introduction

Doxycycline post-exposure prophylaxis (doxy-PEP) is highly effective at reducing bacterial sexually transmitted infections (STIs) among men who have sex with men and transgender women (MSMTW).^1–3^ Implementation of doxy-PEP by local United States health departments began as early as the fall of 2022,^4^ with more widespread uptake after the Centers for Disease Control and Prevention recommended its use in June 2024.^5^ Over the past several years, doxy-PEP has led to population-level reductions in syphilis and chlamydia incidence in MSMTW, with mixed effects on gonorrhea incidence.^6–12^ However, doxy-PEP use has also been associated with increased tetracycline resistance in *Neisseria gonorrhoeae* and *Staphylococcus aureus*.^13–15^ The potential for doxy-PEP to drive antimicrobial resistance in these and other pathogens remains a major concern of both prescribers and patients.^16–18^

The extent to which doxy-PEP drives population-level increases in antimicrobial resistance will depend in part on how much its use increases or decreases net antibiotic use. While doxy-PEP is itself an antibiotic, doxy-PEP recipients can expect reduced antibiotic exposure for STI treatment as a direct effect of doxy-PEP initiation. Furthermore, people in the sexual networks of doxy-PEP recipients who are not taking doxy-PEP themselves may receive fewer antibiotic prescriptions as an indirect effect of doxy-PEP-driven decreases in circulating bacterial STIs.

Prior work estimating the direct effect of doxy-PEP initiation on antibiotic use suggests that initiation of doxy-PEP will drive up net antibiotic use despite reduced prescriptions for STIs.^19^ However, a recent study found that patients taking doxy-PEP who were at high risk for bacterial STIs were exposed to antibiotics for fewer days than patients who were not taking doxy-PEP.^20^ The indirect effects of doxy-PEP have not yet been characterized, but recent work has demonstrated decreases in syphilis incidence in populations that are not currently doxy-PEP eligible after doxy-PEP implementation.^21^ The net impact of doxy-PEP on population-level antibiotic use and antimicrobial resistance will depend on the interplay of these direct and indirect effects.

Here, we retrospectively assessed how doxy-PEP implementation at a sexual health clinic in Boston, Massachusetts changed the number of antibiotic prescriptions received by a cohort of doxy-PEP-eligible MSMTW. The rollout of doxy-PEP at this clinic was associated with significant decreases in diagnoses of chlamydia, which is preferentially treated with doxycycline, and syphilis, which is preferentially treated with penicillin, but not gonorrhea, which is preferentially treated with ceftriaxone.^8^ We aimed to determine 1) the direct effect of current doxy-PEP use on the mean number of antibiotic prescriptions among doxy-PEP users; 2) the indirect effect of cumulative doxy-PEP starts on the mean number of non-doxy-PEP antibiotic prescriptions in people who did and did not start doxy-PEP; 3) the combined impact of the direct and indirect effects on antibiotic prescriptions among doxy-PEP users, doxy-PEP non-users, and the total clinic population.

## Methods

### Study Site

The Massachusetts General Hospital (MGH) Sexual Health Clinic (SHC) is an urban clinic in Boston, MA that provides HIV pre-exposure prophylaxis (PrEP) and STI prevention, testing, and treatment. SHC providers began offering doxy-PEP to MSMTW with bacterial STI risk factors in spring 2023. Patients who started doxy-PEP were asked to return every three months for STI testing and prescription renewals. Comprehensive information about all patients’ sexual behaviors, diagnoses, and treatment was recorded at each visit in a clinic REDCap database.^22,23^ Use of the database for this study was approved by the Institutional Board (IRB) of Mass General Brigham (IRB number: 2003P000336).

### Cohort

Detailed patient visit data, including antibiotic prescriptions and dates of doxy-PEP initiation, were available in the SHC REDCap database from July 1, 2019, until May 14, 2026. Our cohort included all patients who were assigned male at birth, were ≥ 18 years of age, reported sexual partners who identified as men, received HIV PrEP from the clinic after July 1, 2022, and had clinic visits between September 1, 2022, and December 27, 2025. Patients were considered part of the cohort from their first clinic visit until four months after their last clinic visit, in line with the anticipated visit schedule. Prescription rates were assessed between January 1, 2023, and December 27, 2025. We chose to limit this analysis to HIV PrEP users to identify a group of patients who consistently received care at the SHC. Our database only indicated if patients’ sexual partners identified as men, women, or transgender/nonbinary without information about partners’ sex assigned at birth. Patients who reported only transgender/nonbinary sexual partners were included in a sensitivity analysis.

### Variable Definitions

In the SHC clinic database, prescriptions for doxy-PEP were captured separately from prescriptions for non-doxy-PEP doxycycline. Patients were defined as active doxy-PEP users from the date of their first doxy-PEP prescription until the date that they left the clinic cohort. Patients who did not receive any doxy-PEP prescriptions during the period of interest were defined as doxy-PEP non-users, while patients who received at least one doxy-PEP prescription at any point during the study period were defined as doxy-PEP users. This effectively split our population into three groups: patients who were actively taking doxy-PEP, patients who were not yet taking doxy-PEP but eventually started it during the study period, and patients who did not start taking doxy-PEP during the study period. Antibiotic prescriptions in each group were aggregated by week to reduce the impact of weekends and holidays on antibiotic counts. If patients received prescriptions for bacterial STI treatment on the same date that they received their first prescription for doxy-PEP, the STI treatment prescriptions were counted as part of the pre-doxy-PEP period, and the doxy-PEP prescription was counted as part of the post-doxy-PEP period. Each antibiotic prescription was considered equivalent, regardless of the number of doses prescribed or the expected half-life of the medication. We opted to measure prescriptions rather than days of antibiotic exposure due to a lack of data about how patients used doxy-PEP.

### Model

We used Bayesian negative binomial regression implemented in INLA^24,25^ to assess the impact of doxy-PEP implementation on the mean number of prescriptions per 1000 patients. Our covariates of interest were current use of doxy-PEP, which was intended to capture the “direct” effect of doxy-PEP, and the number of people taking doxy-PEP in the total clinic population, which was intended to capture the “indirect” effect of doxy-PEP. We adjusted for baseline differences between doxy-PEP users and doxy-PEP non-users. Among people who were actively using doxy-PEP, we also adjusted for the proportion of patients who had newly started doxy-PEP and by definition received an antibiotic prescription in a given week. Secular time trends were modeled as a first-order random walk to address the effects of temporal autocorrelation. All other effects were modeled as fixed effects. Our main outcome of interest was the mean weekly number of antibiotic prescriptions per 1000 patients in each group, including prescriptions for doxy-PEP. We also performed sub-analyses of the mean weekly number of antibiotic prescriptions per 1000 patients for non-doxy-PEP doxycycline (first-line treatment for chlamydia and second-line treatment for syphilis), ceftriaxone (first-line treatment for gonorrhea), and penicillin (first-line treatment for syphilis). Throughout the text, we report the posterior median estimate of the mean prescription rate. Additional details of the regression models are included in the supplementary appendix.

### Statistical Analysis

#### Estimating the Direct and Indirect Effects of Doxy-PEP

To evaluate the direct effects of doxy-PEP, we estimated the effect of current doxy-PEP use among all the patients in our cohort after controlling for the cumulative doxy-PEP starts in the clinic, baseline differences between doxy-PEP users and non-users, the proportion of new doxy-PEP starts in a given week, and secular time trends. To evaluate the indirect effects of doxy-PEP, we estimated the effect of cumulative doxy-PEP starts in the clinic in all the patients in our cohort after controlling for the effect of active doxy-PEP use, baseline differences between doxy-PEP users and non-users, and secular time trends.

#### Evaluating the Combined Impact of the Direct and Indirect Effects

To evaluate the combined impact of the direct and indirect effects, we first separated patients into doxy-PEP users and doxy-PEP non-users. In each population, we modeled a predicted scenario where doxy-PEP was not introduced in the clinic. In doxy-PEP users, we modeled this scenario by setting both the direct and indirect effects to zero. Among doxy-PEP non-users, who by definition were not impacted by the direct effects of doxy-PEP, we modeled this scenario by setting the indirect effect to zero. We then compared these predicted scenarios to smoothed estimates of observed mean prescriptions per 1000 patients per week in doxy-PEP users and doxy-PEP non-users, obtained by modeling the secular time trend as a random walk. To evaluate the effect of the combined direct and indirect effects of doxy-PEP on the full cohort, we averaged the observed total mean antibiotic prescriptions per 1000 patients per week in doxy-PEP users and doxy-PEP non-users, weighted by the number of patients in each group. We then compared the average rates of antibiotic use in the first and last weeks of the study period.

#### Statistical Software

We performed all statistical analyses and data visualization in R 4.5.2.^26^ For data visualization, we used ggplot2^27^ and patchwork.^28^

## Results

### Patient Characteristics

We identified 1460 patients who were ≥ 18 years of age, identified as MSMTW, and received HIV PrEP from the SHC between September 1, 2022, and December 27, 2025. The number of patients meeting these criteria who were followed by the clinic each week varied over time (**Supplemental Figure 1**), with a minimum of 654 patients followed per week and a maximum of 826 patients followed per week. Of the 1460 total patients, 972 started doxy-PEP, and 488 did not. The total number of patients receiving doxy-PEP from the SHC varied over time, rising rapidly from early 2023 until mid-2024 and reaching a plateau of more than 600 patients by early 2025 (**Figure 1**). Patients who started doxy-PEP had more clinic visits and were more likely to have a history of a bacterial STI than patients who did not start doxy-PEP. Age, gender identity, race, and ethnicity were similar between the groups (**Table 1**).

**Figure 1:**
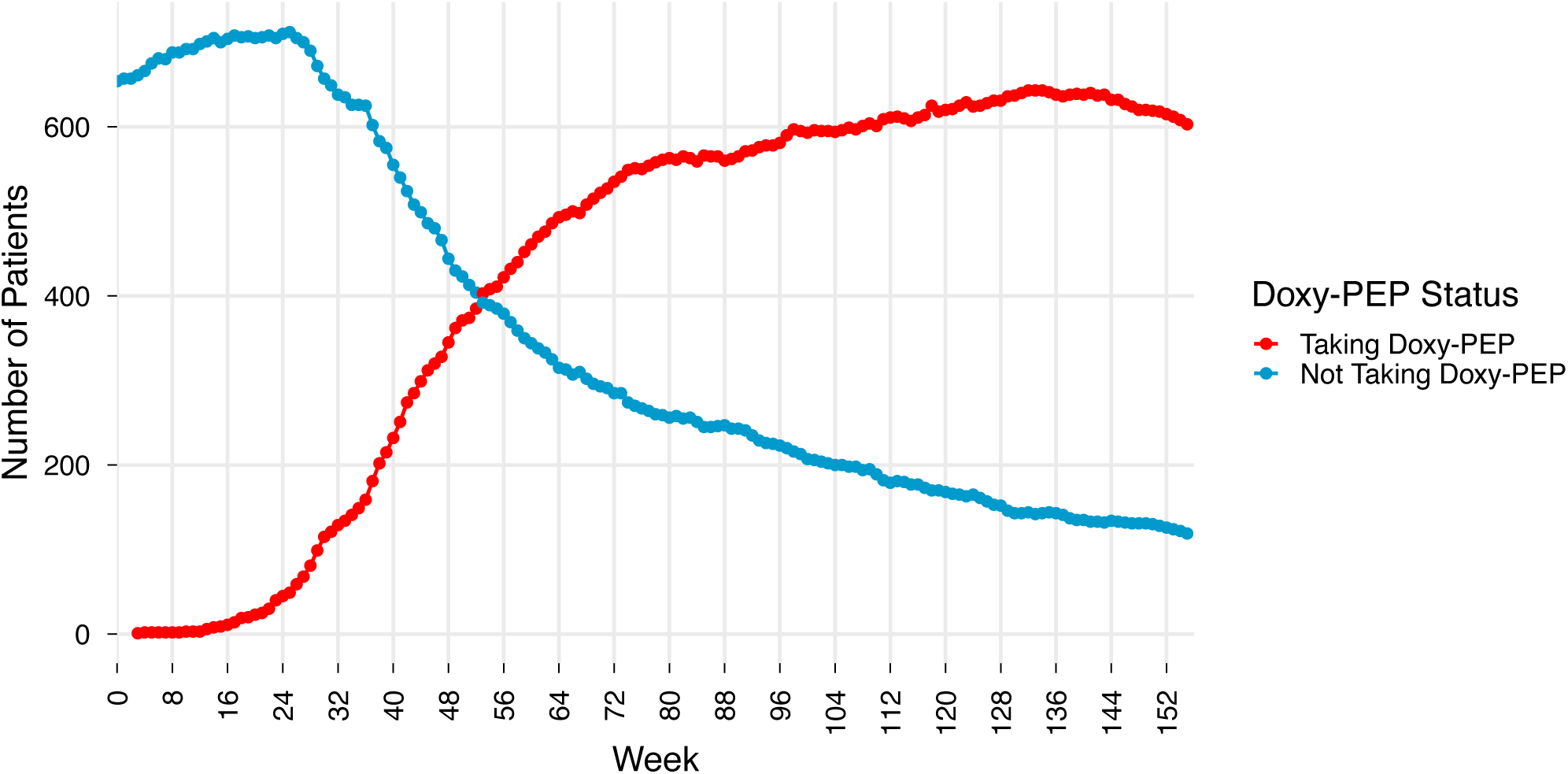
Number of Patients Taking vs Not Taking Doxy-PEP, 2023 – 2025. The number of patients who were included in our cohort and taking or not taking doxy-PEP between January 1, 2023, and December 27, 2025.

**Table 1:**
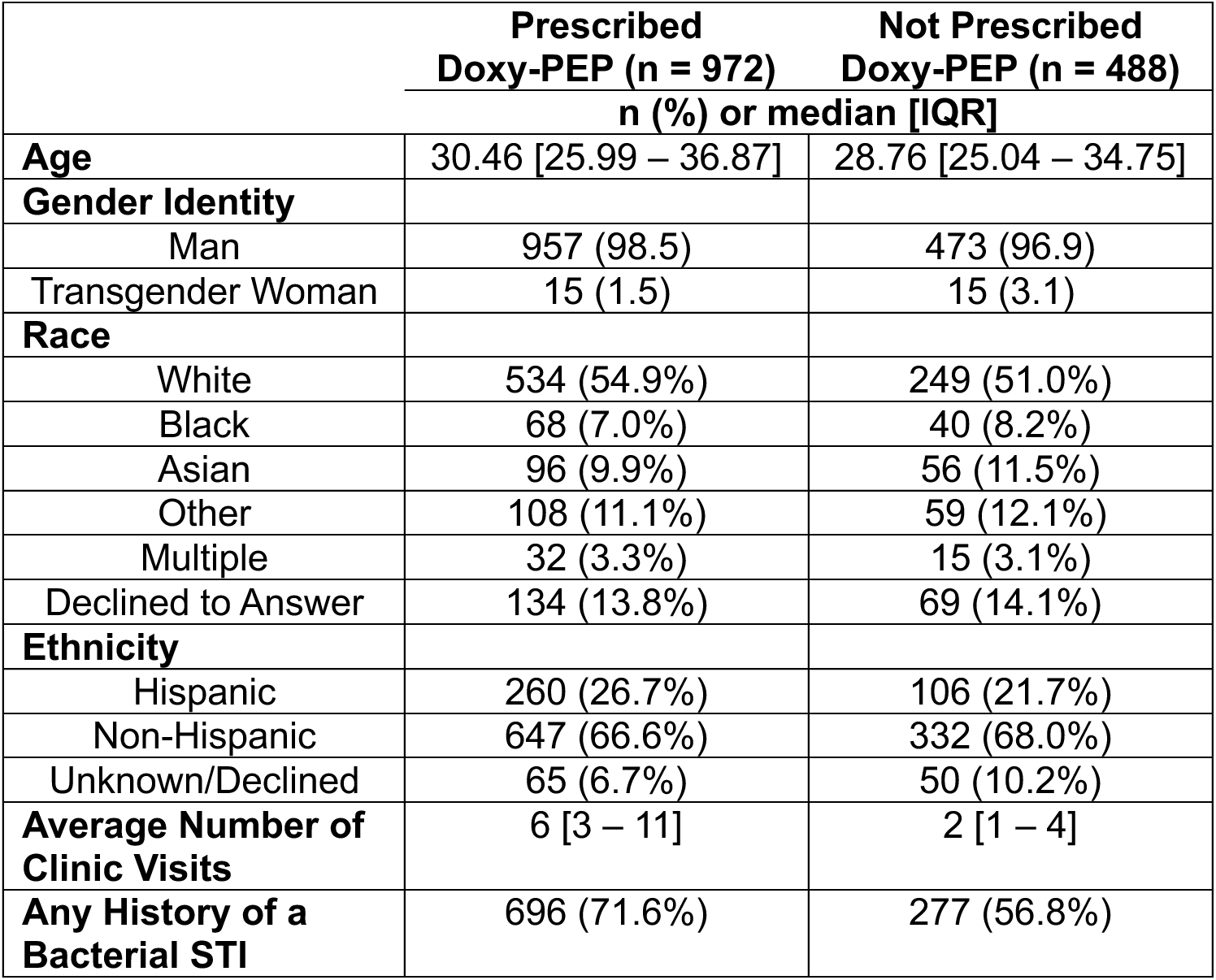
Demographics of MSMTW Patients Taking HIV PrEP Who Did or Did Not Start Doxy-PEP.

### Estimating the Direct and Indirect Effects of Doxy-PEP

The total, non-doxy-PEP doxycycline, ceftriaxone, and penicillin prescriptions per 1000 patients per week in patients who eventually received doxy-PEP and patients who did not varied over the course of the study period (**Supplemental Figure 2**). To assess the direct effect of doxy-PEP, we estimated the effect of current doxy-PEP use on mean total, non-doxy-PEP doxycycline, and ceftriaxone prescriptions per 1000 patients per week. To assess the indirect effect of doxy-PEP, we estimated the effect of cumulative doxy-PEP starts on mean total, non-doxy-PEP doxycycline, and ceftriaxone prescriptions per 1000 patients (**Table 2**). Because of intermittent penicillin shortages throughout the study period, there was insufficient data to draw conclusions about the impact of doxy-PEP on penicillin prescriptions.

**Table 2:** Median Fold Change in Antibiotic Prescriptions (Rx) Per 1000 Patients Per Week Attributable to Doxy-PEP.

|  | Change in Antibiotic Rx Per 1000 Patients Per Week After Starting Doxy-PEP (Direct Effect) | Change in Antibiotic Rx Per 1000 Patients Per Week per 100 Doxy-PEP Starts in the Clinic (Indirect Effect) |
| --- | --- | --- |
|  | Median Fold Change (95% CI) |  |
| Total Antibiotic Rx | 3.40 (2.95 – 3.91) | 0.90 (0.87 – 0.94) |
| Non-Doxy-PEP Doxycycline Rx | 0.37 (0.27 – 0.49) | 0.93 (0.88 – 0.99) |
| Ceftriaxone Rx | 0.77 (0.61 – 0.97) | 0.95 (0.90 – 1.01) |

After controlling for the cumulative doxy-PEP starts in the clinic, the proportion of patients on doxy-PEP who had newly started doxy-PEP and therefore received a prescription in a given week, and differences in the baseline characteristics of the patients who did and did not start doxy-PEP, patients currently using doxy-PEP received 3.40 (95% CI 2.95 – 3.91) times the prescriptions of those not currently using doxy-PEP. Patients currently using doxy-PEP also received 63% fewer non-doxy-PEP doxycycline prescriptions (0.37, 95% CI 0.27 – 0.49) and 23% fewer ceftriaxone prescriptions (0.77, 95% CI 0.61 – 0.97) compared to patients who were not currently using doxy-PEP.

After controlling for the effect of current doxy-PEP use, the proportion of patients on doxy-PEP who had newly started doxy-PEP and therefore received a prescription in a given week, and differences in the baseline characteristics of the patients who did and did not start doxy-PEP, both doxy-PEP users and non-users received 10% fewer mean total antibiotic prescriptions (0.90, 95% CI 0.87 – 0.94) for every 100 doxy-PEP starts in the clinic. There was also a 7% decrease in mean non-doxy-PEP doxycycline prescriptions (0.93, 95% CI 0.88 – 0.99) for every 100 doxy-PEP starts in the clinic. The indirect effect of doxy-PEP implementation on ceftriaxone (0.95, 95% CI 0.90 – 1.01) was not significant.

### Evaluating the Combined Impact of the Direct and Indirect Effects

To evaluate the combined impact of the direct and indirect effects, we separated doxy-PEP users from doxy-PEP non-users and then compared predicted mean antibiotic prescriptions per 1000 patients per week if doxy-PEP had not been introduced to observed antibiotic prescriptions per 1000 patients per week (**Figure 2, Supplemental Table 1**). Among doxy-PEP users, there was a nearly twofold increase in the observed total antibiotic prescription rate compared to the predicted rate, with a median of 35.1 (95% CI 28.4 – 43.5) observed and 18.5 (95% CI 13.8 – 23.1) predicted mean total prescriptions per 1000 patients in the final week of our analysis (**Figure 2a**). The observed mean non-doxy-PEP doxycycline prescription rate was less than one-third the predicted rate, with a median of 1.8 (95% CI 1.2 – 2.7) observed and 6.0 (95% CI 4.4 – 8.0) predicted mean prescriptions per 1000 patients in the final week of our analysis (**Figure 2c**). The observed mean ceftriaxone prescription rate was less than the predicted rate, with a median of 4.9 (95% CI 3.8 – 6.1) observed and 7.5 (95% CI 5.7 – 10.4) predicted mean prescriptions per 1000 patients in the final week of our analysis (**Figure 2e**).

**Figure 2:**
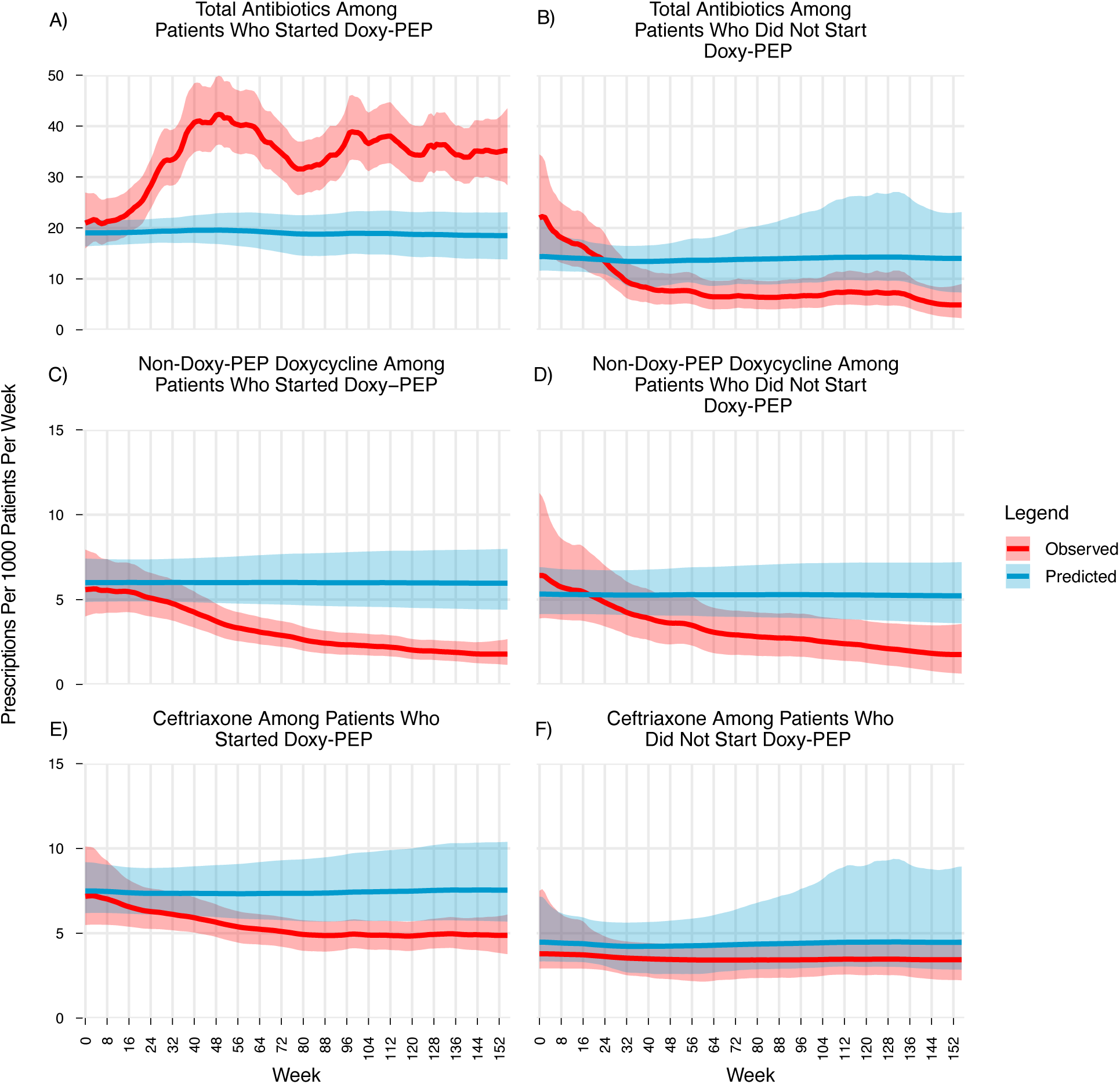
Predicted vs Observed Antibiotic Prescriptions Per 1000 Patients Per Week Among Patients Who Did or Did Not Start Doxy-PEP, 2023-2025. The predicted and observed mean total antibiotic prescriptions including doxy-PEP per 1000 patients per week among A) people who started or B) did not start doxy-PEP; the predicted and observed mean non-doxy-PEP doxycycline prescriptions per 1000 patients per week among C) people who started and D) people who did not start doxy-PEP; and the predicted and observed mean ceftriaxone prescriptions per 1000 patients per week among E) people who started and F) people who did not start doxy-PEP between January 1, 2023 and December 27, 2025. Observed and predicted prescription rates were modeled using Bayesian negative binomial regression implemented in INLA^24,25^ with secular time trends modeled as a random walk. Bold lines represent the predicted (blue) and observed (red) posterior medians, and the shading represents 95% credible intervals.

Among doxy-PEP non-users, the observed mean total antibiotic prescription rate was less than half the predicted prescription rate, with a median of 4.9 (95% CI 2.3 – 9.0) observed and 14.0 (95% CI 7.3 – 23.1) predicted prescriptions per 1000 patients in the final week of our analysis (**Figure 2b**). The observed mean non-doxy-PEP doxycycline prescription rate was a third of the predicted rate, with a median of 1.8 (95% CI 0.64 – 3.6) observed and 5.2 (95% CI 3.6 – 7.2) predicted mean prescriptions per 1000 patients in the final week of our analysis (**Figure 2d**). Given considerable overlap in the credible intervals between the observed and predicted prescription rates for ceftriaxone in the patients who did not start doxy-PEP, a clear effect could not be ascertained (**Figure 2f**).

To evaluate how doxy-PEP implementation changed the mean number of antibiotic prescriptions per 1000 patients per week in our full cohort, we averaged the modeled observed prescription rate in doxy-PEP users and doxy-PEP non-users and compared this averaged rate in the first and last weeks of our analysis. In the first week of our analysis, our cohort included 654 MSMTW taking HIV PrEP, of whom 414 eventually started doxy-PEP and 240 did not start doxy-PEP. Patients in the group who eventually started doxy-PEP received a mean estimate of 21.0 (95% CI 16.0 – 26.9) prescriptions per 1000 patients, and patients who did not eventually start doxy-PEP received a mean estimate of 22.0 (95% CI 14.6 – 34.4) total prescriptions per 1000 patients, with a weighted average of 21.4 total prescriptions per 1000 patients. In the last week of our analysis, our cohort included 722 MSMTW taking HIV PrEP, of whom 603 were taking doxy-PEP and 119 were not. Patients who started doxy-PEP received a mean estimate of 35.1 (95% CI 28.4 – 43.5) total prescriptions per 1000 patients, and patients who did not start doxy-PEP received a mean estimate of 4.9 (95% CI 2.3 – 9.0) total prescriptions per 1000 patients, with a weighted average of 30.1 total prescriptions per 1000 patients. From the beginning of the study period to the end of the study period, the proportion of patients in our cohort who eventually started doxy-PEP increased from 63.3% to 83.5%, and total antibiotic prescribing increased by 1.4 times (**Supplemental Table 2**).

As a sensitivity analysis, we estimated the direct, indirect, and combined effects of doxy-PEP on a cohort that included patients whose only sexual partners identified as transgender/nonbinary with unspecified sex assigned at birth. There were no substantive changes to our findings (**Supplemental Table 3**).

## Discussion

We evaluated how implementation of doxy-PEP at a Boston sexual health clinic changed the number of antibiotic prescriptions received by a cohort of MSMTW who were taking HIV PrEP. We estimated the impact of current doxy-PEP use on prescription rates to capture the “direct” effect of doxy-PEP and the impact of cumulative doxy-PEP starts in the clinic on prescription rates to capture the “indirect” effect of doxy-PEP. We then evaluated how these effects combined to change mean antibiotic prescription rates in doxy-PEP users and non-users. Finally, we reported how doxy-PEP implementation changed the total prescription rate in our full cohort between the first and last weeks of the study period.

After controlling for cumulative doxy-PEP starts in the clinic, patients who were actively using doxy-PEP received 3.4 times the prescriptions of patients who were not actively taking doxy-PEP. However, after controlling for active doxy-PEP use, the total antibiotic prescription rate decreased by about 10% for every 100 doxy-PEP starts in the clinic. When we assessed the combined impact of these direct and indirect effects among patients who started doxy-PEP, the observed prescription rate in this group was nearly twice the predicted rate if doxy-PEP had not been implemented. Among patients who did not start doxy-PEP, the observed prescription rate was less than half of the predicted rate if doxy-PEP had not been implemented. When comparing the number of antibiotic prescriptions in the first and last weeks of our analysis in our full cohort of doxy-PEP users and doxy-PEP non-users, doxy-PEP implementation was associated with a 1.4-fold increase in antibiotic prescribing.

Doxy-PEP directly and indirectly decreased the prescription rate for non-doxy-PEP doxycycline, reflecting reductions in chlamydia and syphilis that had previously been reported at the SHC after doxy-PEP implementation.^8^ Despite prior findings that there was not a significant decrease in total gonorrhea diagnoses after doxy-PEP implementation, we also found that doxy-PEP directly decreased the ceftriaxone prescription rate. Potential explanations for this discrepancy are the inclusion of nearly two additional years of data in our study as well as fewer cases of symptomatic urethritis requiring empiric treatment with ceftriaxone and doxycycline prior to identification of a causative organism. We had insufficient data to draw conclusions about the direct and indirect effects of doxy-PEP on penicillin prescriptions in the setting of intermittent shortages after April 2023 that prompted conservation of benzathine penicillin for use in pregnant women and infants with syphilis.^29^

Our results differ from a similar analysis of patients followed by a sexual health clinic in Milan, Italy, where doxy-PEP initiation directly reduced days of antibiotic exposure.^20^ This difference may be partially driven by selective initiation of doxy-PEP in patients who were felt to be at particularly high risk for bacterial STIs in the Milan cohort. Prior modeling demonstrated that doxy-PEP is most efficient when prescribed preferentially to the subset of patients most likely to contract STIs.^30^ As the incidence of bacterial STIs decreases, the direct effect of doxy-PEP can be expected to skew toward increased net antibiotic exposure, while the indirect effect will become less prominent.

While our analysis was only performed in MSMTW, bridging by men who have sex with men and women may extend the indirect effects of doxy-PEP to sexual networks that include cisgender women. This is one potential explanation for recent findings that doxy-PEP implementation in Seattle King County was associated with a significant decrease in the incidence of syphilis among cisgender women, a group for whom doxy-PEP is not currently recommended.^21^

This study had several limitations. First, we chose to use antibiotic prescriptions rather than days of antibiotic exposure as our primary outcome due to a lack of consistent data about how patients were taking doxy-PEP. Given that a quarter of patients in the DoxyPEP trial reported one or fewer monthly uses of doxy-PEP,^2^ our choice to use prescriptions as our primary outcome may overestimate antibiotic exposure after initiation of doxy-PEP in infrequent doxy-PEP users. Second, this analysis was performed in one Boston clinic specializing in sexual health care and may not generalize to all populations of patients who take doxy-PEP. Third, we were unable to account for doxy-PEP prescriptions or prescriptions for STI treatment from outside the SHC. Finally, we do not have information about patients’ sexual networks and instead assumed shared sexual networks based on geographic proximity and patient-reported sexual behavior.

In conclusion, even when accounting for decreased instances of STI treatment after initiation of doxy-PEP, MSMTW in our clinic population received nearly twice as many prescriptions as patients not taking doxy-PEP. However, doxy-PEP implementation was associated with a decrease in the antibiotic prescription rate among MSMTW who were not using doxy-PEP.

While individuals who are taking doxy-PEP have higher levels of antibiotic exposure and are therefore at increased risk for infection with antibiotic resistant organisms, doxy-PEP-driven decreases in the incidence of bacterial STIs have the potential to drive down antibiotic use and selection for resistance in populations that are not using doxy-PEP. As the incidence and prevalence of the bacterial STIs targeted by doxy-PEP change, the extent of doxy-PEP’s direct and indirect effects should also be expected to vary and will need continued study. Future studies are also needed to evaluate the direct and indirect effects of doxy-PEP on population-level antibiotic resistance.

## Data Availability

To maintain patient confidentiality, we cannot share disaggregated data from electronic health records. All code is available on GitHub at https://github.com/gradlab/SHC_Doxy_PEP_Antibiotic_Use.

