## Supplemental Material for "The Impact of Doxycycline Post-Exposure Prophylaxis on Antibiotic Use at a Boston Sexual Health Clinic"

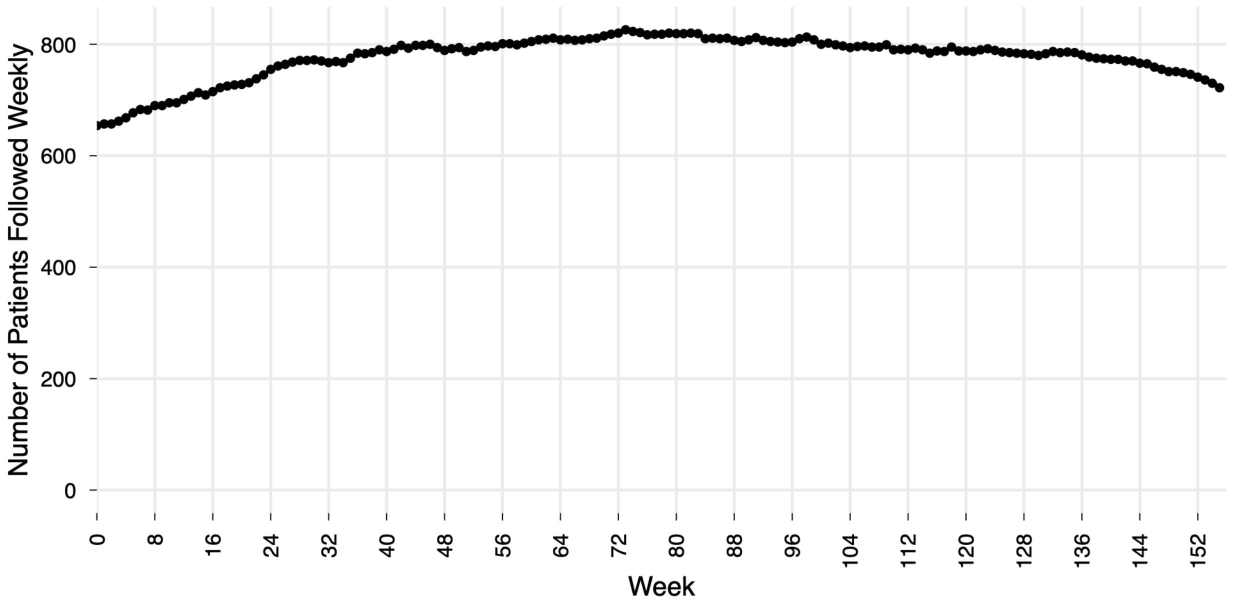

**Supplemental Figure 1: Number of MSMTW Taking HIV-PrEP Followed by the Sexual Health Clinic 2023- 2025.** Patients were considered part of the cohort from their first clinic visit until four months after their last clinic visit. Visit data spanned July 1, 2019, to May 14, 2026. For this analysis, prescription rates were evaluated between January 1, 2023, which corresponds with week 0 above, and December 27, 2025, which corresponds with week 155 above. MSMTW = men who have sex with men and trans women; HIV-PrEP = HIV pre-exposure prophylaxis.

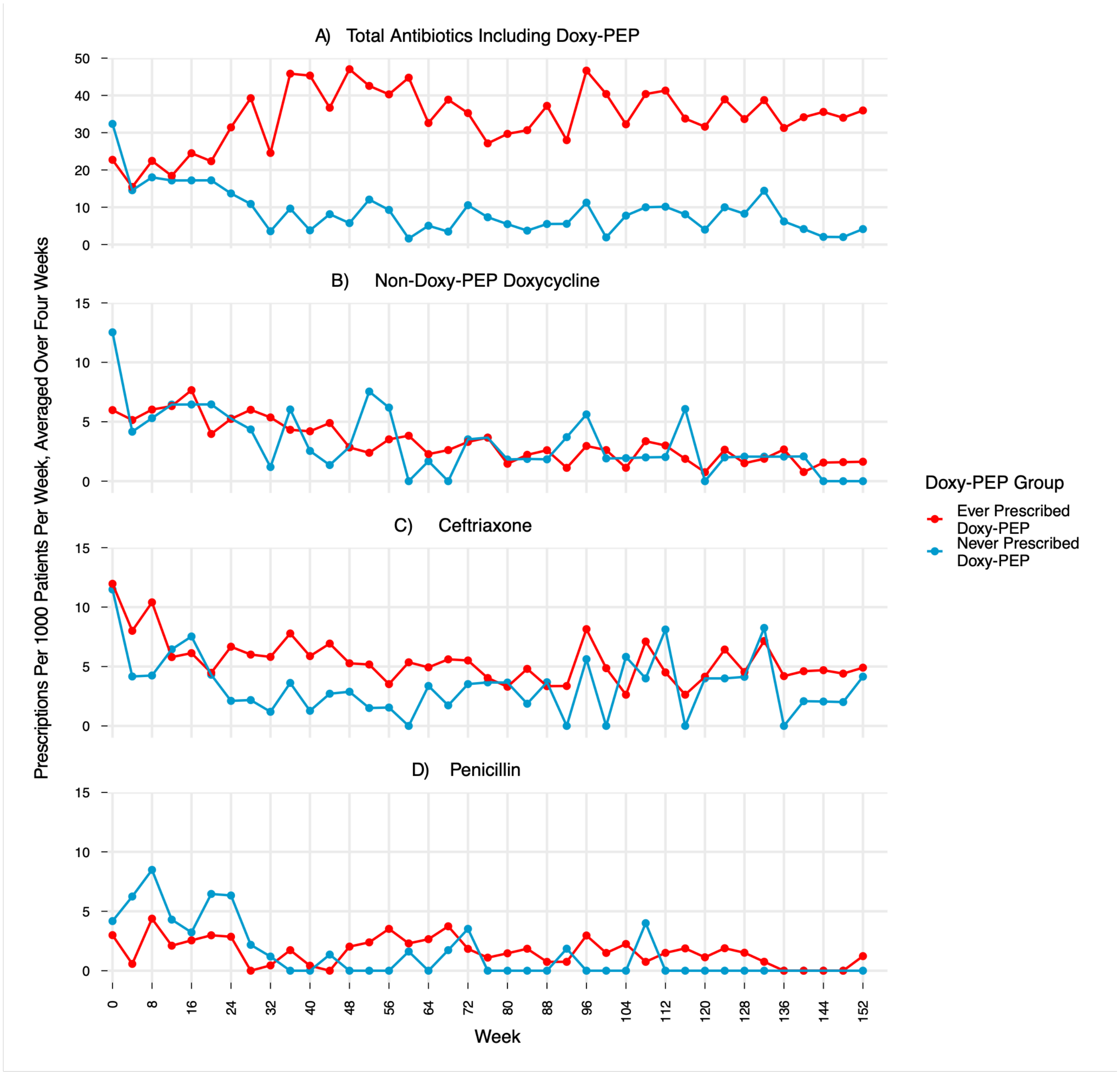

**Supplemental Figure 2: Observed Antibiotic Prescriptions Per 1000 Patients Per Week Among Patients Who Did or Did Not Start Doxy-PEP.** Total antibiotic prescriptions including doxy-PEP (A), non-doxy-PEP doxycycline prescriptions (B), ceftriaxone prescriptions (C), and penicillin prescriptions (D) per 1000 patients per week in patients who eventually started (red) or never started (blue) doxy-PEP. Prescriptions are averaged over 4 weeks for ease of interpretation, with points at the first week of each 4-week block.

**Supplemental Table 1: Observed versus Predicted Mean Prescriptions per 1000 Patients per Week in the Final Week of the Study Period**

| | Observed Mean Prescriptions per 1000 Patients per Week (95% CI) | Predicted Mean Prescriptions per 1000 Patients per Week (95% CI) |
|---|---|---|
| **Total Antibiotics** | | |
| Doxy-PEP Users | 35.1 (28.4 – 43.5) | 18.5 (13.8 – 23.1) |
| Doxy-PEP Non-Users | 4.9 (2.3 – 9.0) | 14.0 (7.3 – 23.1) |
| **Non-Doxy-PEP Doxycycline** | | |
| Doxy-PEP Users | 1.8 (1.2 – 2.7) | 6.0 (4.4 – 8.0) |
| Doxy-PEP Non-Users | 1.8 (0.64 – 3.6) | 5.2 (3.6 – 7.2) |
| **Ceftriaxone** | | |
| Doxy-PEP Users | 4.9 (3.8 – 6.1) | 7.5 (5.7 – 10.4) |
| Doxy-PEP Non-Users | 3.4 (2.2 – 4.6) | 4.5 (2.9 – 8.9) |

**Supplemental Table 2: Weighted Average Antibiotic Prescriptions per Week in the Full Cohort in the First and Last Weeks of the Study Period**

| | Patient Count | Total Antibiotic Prescriptions per 1000 Patients per Week |
|---|---|---|
| **Week 1** | | |
| Doxy-PEP Users | 414 | 21.0 (95% CI 16.0 – 26.9) |
| Doxy-PEP Non-Users | 240 | 22.0 (95% CI 14.6 – 34.4) |
| Weighted Average | | 21.4 |
| **Week 156** | | |
| Doxy-PEP Users | 603 | 35.1 (95% CI 28.4 – 43.5) |
| Doxy-PEP Non-Users | 119 | 4.9 (95% CI 2.3 – 9.0) |
| Weighted Average | | 30.1 |

**Supplemental Table 3: Findings When Including vs Not Including Patients Assigned Male at Birth Who Reported Transgender/Nonbinary Sexual Partners**

| | **Not Including Patients Assigned Male at Birth with Only Transgender or Nonbinary Sexual Partners** | **Including Patients Assigned Male at Birth with Only Transgender or Nonbinary Sexual Partners** |
|---|---|---|
| **Number of Patients** | Count | |
| Total | 1460 | 1475 |
| Doxy-PEP Users | 972 | 979 |
| Doxy-PEP Non-Users | 488 | 496 |
| **Direct Effect** | Median Fold Change (95% CI) | |
| Total | 3.40 (2.95 – 3.91) | 3.44 (2.99 – 3.96) |
| Non-Doxy-PEP Doxycycline | 0.37 (0.27 – 0.49) | 0.37 (0.28 – 0.50) |
| Ceftriaxone | 0.77 (0.61 – 0.97) | 0.78 (0.62 – 0.99) |
| **Indirect Effect** | Median Fold Change (95% CI) | |
| Total | 0.90 (0.87 – 0.94) | 0.90 (0.87 – 0.94) |
| Non-Doxy-PEP Doxycycline | 0.93 (0.88 – 0.99) | 0.93 (0.88 – 0.99) |
| Ceftriaxone | 0.95 (0.90 – 1.01) | 0.95 (0.89 – 1.01) |
| **Combined Effect: Doxy-PEP Users, Final Week of Analysis** | Prescriptions Per 1000 Patients Per Week (95% CI) | |
| Total Observed | 35.1 (28.4 – 43.5) | 35.1 (28.4 – 43.4) |
| Total Predicted | 18.5 (13.8 – 23.1) | 18.4 (13.8 – 22.9) |
| Non-Doxy-PEP Doxycycline Observed | 1.8 (1.2 – 2.7) | 1.8 (1.2 – 2.7) |
| Non-Doxy-PEP Doxycycline Predicted | 6.0 (4.4 – 8.0) | 5.9 (4.4 – 8.0) |
| Ceftriaxone Observed | 4.9 (3.8 – 6.1) | 4.8 (3.7 – 6.1) |
| Ceftriaxone Predicted | 7.5 (5.7 – 10.4) | 7.5 (5.7 – 10.3) |
| **Combined Effect: Doxy-PEP Non-Users, Final Week of Analysis** | Prescriptions Per 1000 Patients Per Week (95% CI) | |
| Total Observed | 4.9 (2.3 – 9.0) | 4.7 (2.1 – 8.8) |
| Total Predicted | 14.0 (7.3 – 23.1) | 14.4 (6.2 – 23.5) |
| Non-Doxy-PEP Doxycycline Observed | 1.8 (0.64 – 3.6) | 1.7 (0.60 – 3.5) |
| Non-Doxy-PEP Doxycycline Predicted | 5.2 (3.6 – 7.2) | 5.3 (3.6 – 7.3) |
| Ceftriaxone Observed | 3.4 (2.2 – 4.6) | 3.5 (2.3 – 4.6) |
| Ceftriaxone Predicted | 4.5 (2.9 – 8.9) | 4.5 (2.9 – 8.1) |
| **Observed Total Antibiotic Use Per 1000 Patients Per Week, Total Cohort** | Average Prescriptions Per 1000 Patients Per Week | |
| First Week | 21.4 | 22.1 |
| Last Week | 30.1 | 30.1 |

### Appendix

#### Model equations

At time $i$ in group $j$, which has $N_{ij}$ total patients, the mean number of prescriptions per patient is denoted by $\mu_{ij}$. We model $\log(\mu_{ij})$ as a linear function of the following covariates: *post_dpep_start*, the effect of starting doxy-PEP, which is 0 in patients who have not started doxy-PEP and 1 in patients who have started it; *dpep_ever*, the effect of being in the group that did or did not start doxy-PEP, which is 0 for patients who never start doxy-PEP and 1 for patients who eventually start it; *prop_new_dpep*, a variable equal to the proportion of patients on doxy-PEP (*post_dpep_start* = 1, *dpep_ever* = 1) who have newly started doxy-PEP in a given week and therefore received a prescription, and zero in all other compartments; and *cumulative_dpep100*, a count of the number of patients who have started doxy-PEP in our cohort divided by 100. The secular time trend $\varepsilon(t_i)$ is modeled as a random walk. The prescription count $y_{ij}$ is modeled using a negative binomial distribution with mean $\mu_{ij}N_{ij}$ and overdispersion parameter $\theta$.

<u>Linear predictor</u>

$$log(\mu_{ij}) \sim \beta_1 post\_dpep\_start_{ij} + \beta_2 dpep\_ever_{ij} + \beta_3 prop\_new\_dpep_{ij} + \beta_4 cumulative\_dpep100_i + \varepsilon(t_i)$$

<u>Mean overdispersion parametrization</u>

$$y_{ij} |\ \mu_{ij}, \theta, N_{ij} \sim NB(\mu_{ij} N_{ij}, \theta)$$
